## Supporting Information for "Population birth outcomes in 2020 and experiences of expectant mothers during the COVID-19 pandemic: a ‘Born in Wales’ mixed methods study using routine data"

**S1 Fig. Coding framework detailing the themes, subthemes and definitions from the qualitative analysis of women’s experiences of pregnancy during the COVID-19 pandemic**

| Theme | Subtheme | Definition |
| --- | --- | --- |
| Perception of the severity of the COVID-19 pandemic | Feeling nervous about the COVID-19 pandemic | Any comments about feeling nervous, worried or anxious about being pregnant during the COVID-19 pandemic; any remarks about the impact of COVID-19 lockdown and other restrictions on social support for expectant parents |
|  | Anxiety about contracting COVID-19 | Any comments about being worried about contracting COVID-19 whilst pregnant, or being afraid to leave the house, go to the shops or attend appointments due to fear of catching the virus |
|  | Impact of COVID-19 and associated restrictions on the health and wellbeing of the baby | Any remarks about the impact of COVID-19-related stress on the unborn child; any comments about the potential impact of stress/the infection itself on foetal development; any remarks about pregnant women and unborn babies as vulnerable groups |
| Difference to regular appointments and delivery | Opinions on virtual appointments and services | Any comments about virtual or telephone appointments/classes/support groups and the impact of virtual vs. face-to-face services on women’s perceived level of support |
|  | Partner’s presence at scans, appointments and delivery and impact on expectant parents mental health | Any remarks about the difficulties of attending scans or appointments alone and the impact of this on the mental health of the expectant mother and partner; any comments about being worried or anxious to give birth alone; respondents’ opinions on the visiting restrictions for partners and the importance of partners attending scans/appointments/delivery |
| Support from midwives | Level of contact and support received from midwives and impact on mental health and enjoyment of pregnancy | Any comments (positive, neutral or negative) about the level of support received from midwives during the COVID-19 pandemic; any remarks about the amount and/or type of contact respondents had received with midwives; any comments about the impact of support/lack of support on the enjoyment of pregnancy or on feelings of anxiety/loneliness/uncertainty; respondents opinions on the level of support offered under such unprecedented circumstances |
|  | Communication issues | Any comments regarding communication from midwives during the COVID-19 pandemic; any comments about feeling uninformed, uncertain or ‘left in the dark’; any remarks about women being unable to access care and support due to lack of communication; any comments about women having to seek out information for themselves online |

**S2 Fig. Software used for analysis**

Data linkage of datasets were performed in SAIL using a combination of  SQL coding in

- Eclipse [1]version 2020-03 (4.15) and;
- in R [2] version 4.0.4, using RStudio [3]version 1.4.1103.

Analysis of the data was also performed in R Studio. Details of the specific software packages[4-12] used within RStudio can be found in the references.

**S3 Fig. SAIL Databank data sources**

(*Data sources from which attributes were selected****)***

|  | **National Community Child Health**  **(NCCH)** | **Primary care**  **Data**  **(WLGP)** | **Secondary care**  **Data**  **(PEDW)** | **Annual District Death Extract**  **(ADDE)** | **Welsh Demographic Service Dataset**  **(WDSD)** |
| --- | --- | --- | --- | --- | --- |
| **Sex** | **✓** |  |  |  |  |
| **Week of birth** | **✓** |  |  |  |  |
| **Birth weight** | **✓** |  |  |  |  |
| **Gestation at birth** | **✓** |  |  |  |  |
| **Stillbirths** | **✓** |  |  |  |  |
| **C-Sections** | **✓** | **✓** | **✓** |  |  |
| **Mortality** | **✓** |  |  | **✓** |  |
| **Deprivation quintiles** |  |  |  |  | **✓** |
| **Rural & Urban classification** |  |  |  |  | **✓** |

**S4 Fig. Data definitions**

| **Stillbirth** | Flagged as stillborn or not in dataset. Definition may vary between units. | | | |
| --- | --- | --- | --- | --- |
| **Gestation at birth**  *(Number of completed weeks of gestation at birth)* | Extremely preterm (EPT) | | | < 28 weeks |
|  | Very preterm (VPT) | | | 28 – 31 weeks |
|  | Preterm (PT) | | | 32 - 36 weeks |
|  | Term (T) | | | 37 – 41 weeks |
|  | Late term (LT) | | | ≥ 42 weeks |
| **Birth weight category** | Extremely Low Birth Weight (ELBW) | | | ≤ 1kg |
|  | Very Low Birth Weight (VLBW) | | | 1.001 - 1.5kg |
|  | Low Birth Weight (LBW) | | | 1.501 - 2.5kg |
|  | Normal Birth Weight (NBW) | | | 2.501 - 4kg |
|  | High Birth Weight (HBW) | | | 4.001 - 4.5kg |
|  | Very High Birth Weight (VHBW) | | | >4.5kg |
| **C-Sections**  *(Elective & Emergency)* | Delivery, International Statistical Classification of Diseases and Related Health Problems , 10^th^ edition (ICD10), and Read v2. Codes from NCCH, PEDW and GP datasets as per codes listed in supplementary data | | | |
| ***Preterm mortality** | Premature neonates (<37 weeks gestation) who do not survive to 40 weeks after the estimated date of conception. | | | |
| ***Neonatal mortality** | All non-preterm babies that do not survive to 28-days after birth. | | | |
| ***Infant mortality** | All non-preterm babies that survive to 28 days after birth but did not survive until 90-days after birth. | | | |
| **Rural/Urban location** | Codes from WDSD 2011 census data. 1 & 2 were combined to represent ‘Urban’ locations; 3 used for ‘Rural’. | | | |
|  | 1 | Town and fringe | | |
|  | 2 | Urban population > 10,000 | | |
|  | 3 | Village, hamlet and isolated dwellings | | |
| **Deprivation** | Welsh Index of Multiple Deprivation (WIMD) version 2019 quintiles: | | | |
|  | Quintile 1 | | – 20% most deprived | |
|  | Quintile 2 | | – 20-40% most deprived | |
|  | Quintiles 3-5 | | – remaining 60% least deprived. | |

********Mortality data for births at the latter end of 2020 were unavailable at the time of analysis beyond 01/02/2021. For births in 2016 to 2019, mortality data extending beyond February 1^st^ of the year post birth were excluded to allow direct comparison with 2020 data.*

1.	Public Health Wales. The Complete Routine Immunisation Schedule from January 2020. 2020. Report No. Available from: <https://111.wales.nhs.uk/pdfs/adultschedule.pdf>.

2.	Public Health Wales. Routine Immunisation Schedule Wales from September 2019. 2019. Report No. Available from: <http://www.wales.nhs.uk/sitesplus/documents/888/Routine%20Immunisation%20Schedule%20Wales%20from%20September%202019.pdf>.

3.	Public Health Wales. Routine Childhood Immunisations from August 2018. 2018. Report No. Available from: <https://www2.nphs.wales.nhs.uk/VaccinationsImmunisationProgsDocs.nsf/3dc04669c9e1eaa880257062003b246b/094ce8b3ed8504b5802582e3002d8600/$FILE/Routine%20Childhood%20Immunisation%20Schedule%20-%207%20August%202018%20Final.pdf>.
